# Assessment of Specimen Pooling to Conserve SARS CoV-2 Testing Resources

**DOI:** 10.1101/2020.04.03.20050195

**Authors:** Baha Abdalhamid, Christopher R. Bilder, Emily L. McCutchen, Steven H. Hinrichs, Scott A. Koepsell, Peter C. Iwen

**Author notes:** **Corresponding Author** Baha Abdalhamid, MD, PhD, D (ABMM), DRC II 7035, Department of Pathology and Microbiology, University of Nebraska Medical Center, 985900 Nebraska Medical Center, Omaha, NE 68198-5900.

## Abstract

**Objectives:** To establish the optimal parameters for group testing of pooled specimens for the detection of SARS-CoV-2.

**Methods:** The most efficient pool size was determined to be 5 specimens using a web-based application. From this analysis, 25 experimental pools were created using 50 microliter from one SARS-CoV-2 positive nasopharyngeal specimen mixed with 4 negative patient specimens (50 microliter each) for a total volume of 250 microliter l. Viral RNA was subsequently extracted from each pool and tested using the CDC SARS-CoV-2 RT-PCR assay. Positive pools were consequently split into individual specimens and tested by extraction and PCR. This method was also tested on an unselected group of 60 nasopharyngeal specimens grouped into 12-pools.

**Results:** All 25 pools were positive with Cycle threshold (Ct) values within 0 and 5.03 Ct of the original individual specimens. The analysis of 60 specimens determined that two pools were positive followed by identification of two individual specimens among the 60 tested. This testing was accomplished while using 22 extractions/PCR tests, a savings of 38 reactions.

**Conclusions:** When the incidence rate of SARS-CoV-2 infection is 10% or less, group testing will result in the saving of reagents and personnel time with an overall increase in testing capability of at least 69%.

**Key Points:** SARS CoV-2, COVID-19, Group testing

---

Since the first detection in Wuhan, China in December 2019, severe acute respiratory syndrome coronavirus 2 (SARS-CoV-2), the pathogen of coronavirus diseases 2019 (COVID-19), has spread worldwide to now be considered a pandemic ^1,2^. The United States (US) is experiencing an acute shortage of certain reagents important for performance of assays for the detection of SARS-CoV-2. Some areas of the US have stopped testing due to lack of test supplies. The ability to rapidly diagnosis COVID-19 is important for evaluating the spread of disease and for tracing the contacts of infected individuals.

The assay developed by the Centers for Disease Control and Prevention (CDC) for detection of SARS-CoV-2 and approved for use under emergency use authorization (EUA) by the FDA has been widely employed by public health laboratories throughout the US^3^. This assay employs an extraction procedure of viral RNA from specimens collected by nasopharyngeal (NP) swabs. The second step in the assay employs reverse transcription and amplification using a real time PCR instrumentation. The assay therefore requires two kits, one for extraction and another for amplification of the target and detection. We investigated whether a strategy used in the testing of blood prior to transfusion could have application for conservation of scarce reagents for the SARS CoV-2 assay ^4,5^. The process of group testing that employs sample pooling is used for detection of the human immunodeficiency virus, and hepatitis B and C viruses^4^ in blood products. Key principles for successful application of group testing involve knowledge of the limit-of-detection, sensitivity and specificity of the assay, and the prevalence of disease in the population. The goal of the process is to determine a pool size that provides the greatest conservation of resources while maintaining the reliable performance of testing. This report describes a proof-of-concept for group testing of pooled specimens for the diagnosis of COVID-19.

To assess the group testing strategy, the first step was to calculate the most efficient pool size using a web-based application for pooling as described at https://www.chrisbilder.com/shiny. Although the prevalence of COVID-19 in Nebraska has not been specifically defined by comprehensive epidemiology studies, the observed specimen positive rate within the tested community has been around 5% for the past two weeks. The following parameters and assumptions used in this calculation included an experimental prevalence rate of 5%, an assay lower limit-of-detection of 1 to 3 RNA copies/µl, an assay sensitivity of 95% or 100%, an assay specificity of 100%, a two-stage pooling algorithm, and a range of pool sizes of 3 to 10 samples^6^. These calculations predicted a pool size of 5 samples would provide the largest reduction in the expected number of tests of 57% when compared to testing specimen separately (Figure 1).

**Figure 1.**
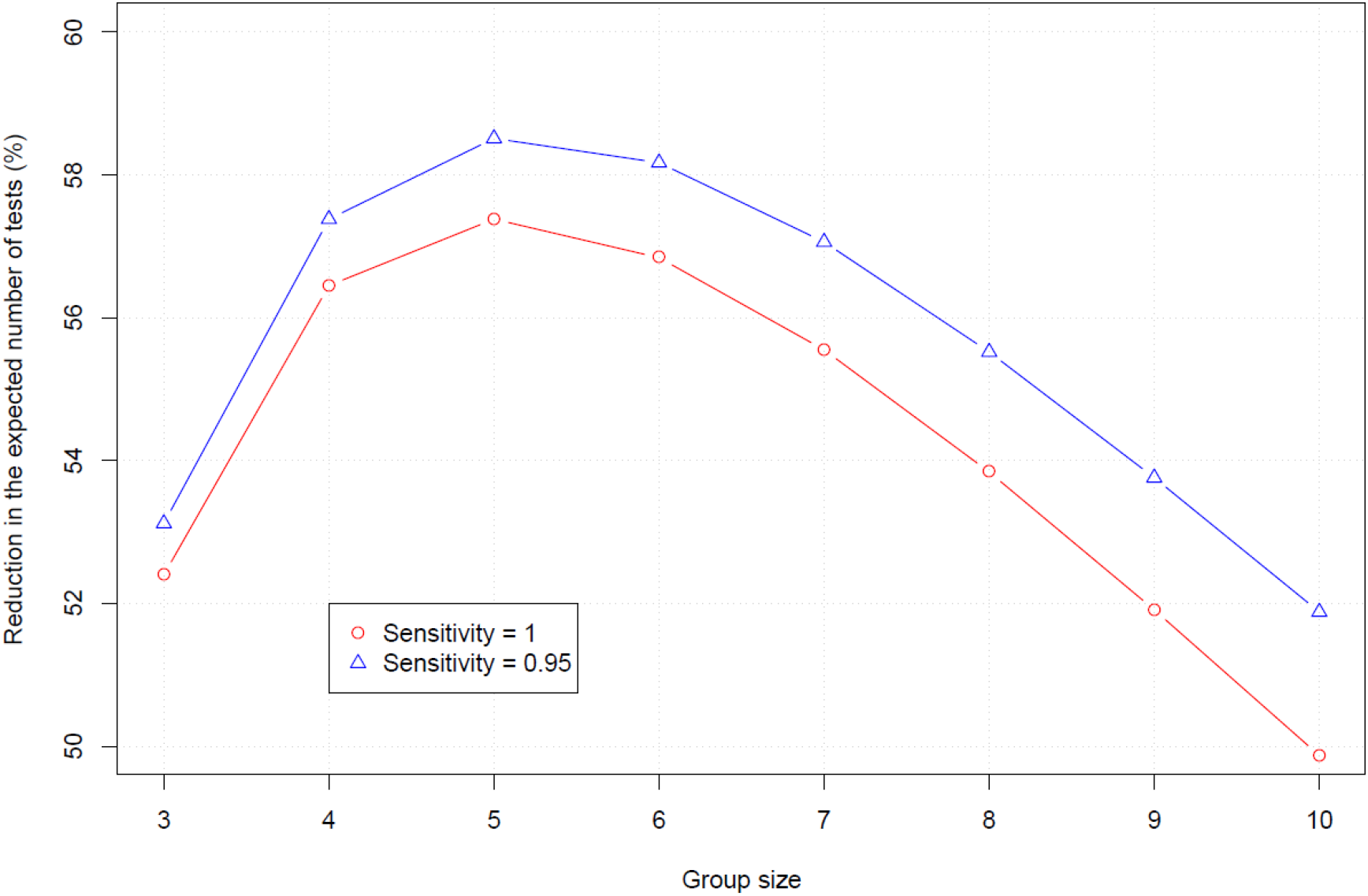
Optimal sample pool size Graphical comparison of initial pool size compared to expected number of tests per individual using the Shiny application for pooled testing available at https://www.chrisbilder.com/shiny. The optimal sample pool size was determined based on the least number of tests and the following parameters: prevalence rate (5%), a lower limit of detection of 1 to 3 RNA copies/µl, an assay sensitivity of either 95% or 100%, and an assay specificity of 100%.

The CDC RT-PCR assay was used in this study to detect SARS-CoV-2 in nasopharyngeal specimens. With this assay, a positive COVID-19 result is determined when both nucleocapsid targets (N1 andN2) reach a defined cycle threshold (Ct) of <40. For 158 confirmed positive specimens that have been seen in the public health laboratory to date, the Ct values for N1 have averaged 26.06 with a SD of 5.5 (range 15.75 to 37.96) and Ct values for N2 have averaged 26.48 with a SD of 5.8 (range 15.75 to 38.65). Twenty-five pools of five specimens with each containing one positive patient were group tested for this study. Of these, the COVID-19 positive specimens were within a range of Ct values from 18.23 to 36.74 for N1 and from 17.33 to 37.43 for N2. Included in this evaluation 14 specimens were selected with low RNA concentration (Ct > 30) (Table 1). Note that a low Ct values indicated the presence of higher amounts of viral RNA and high Ct values indicated lower amounts.

**Table 1.**
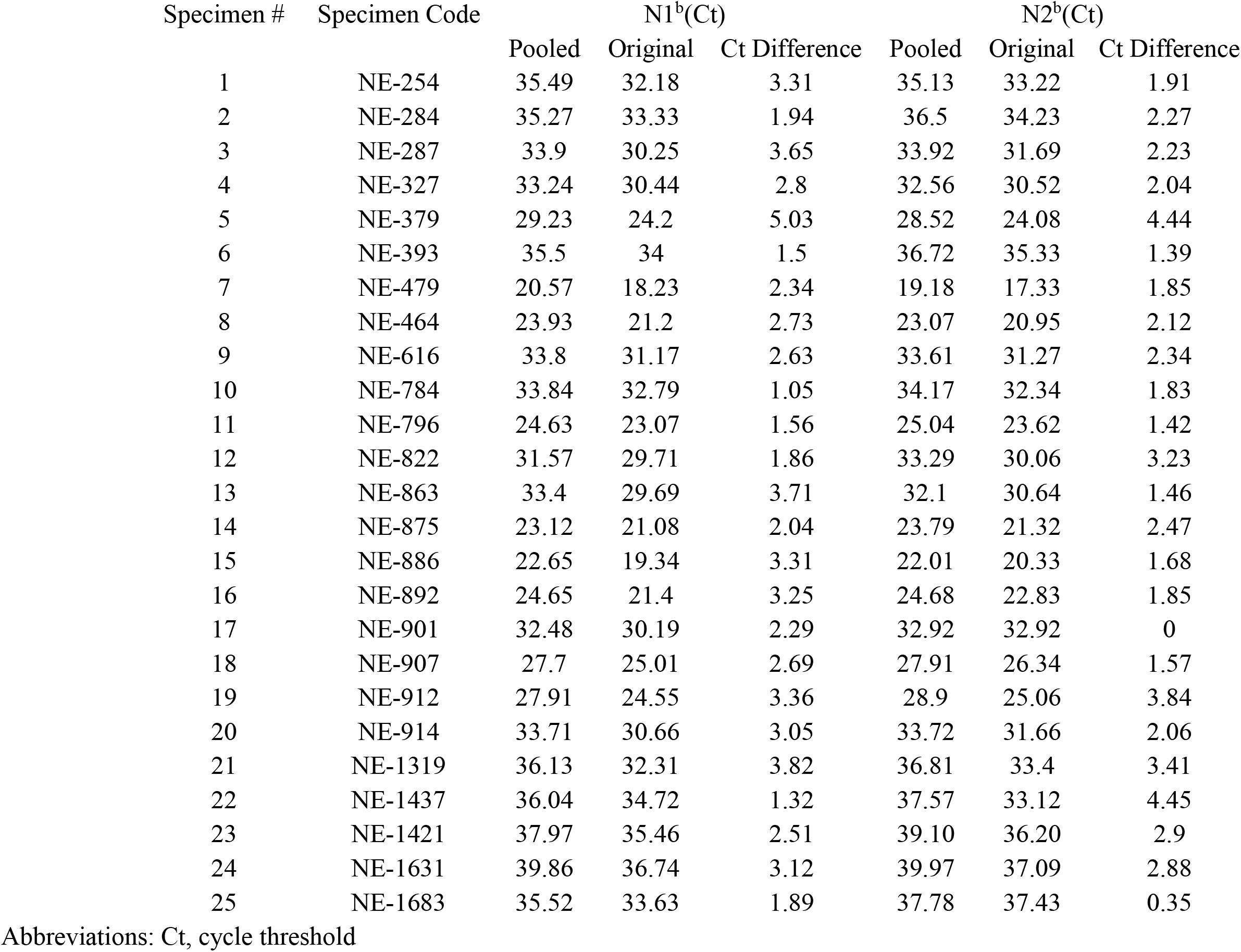
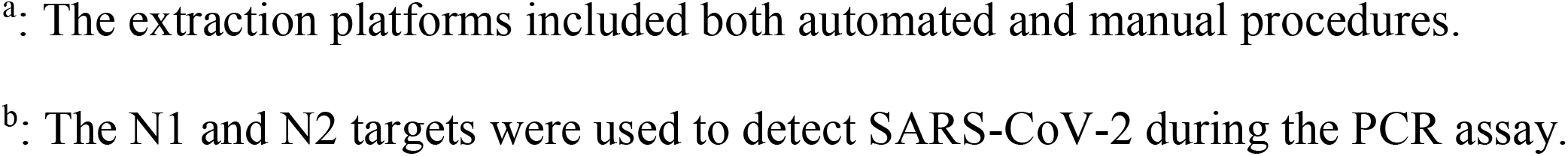
Comparison of threshold cycles between the original and pooled COVID-19 positive samples ^a^.

| Specimen # | Specimen Code | N1<sup>b</sup>(Ct) |  |  | N2<sup>b</sup>(Ct) |  |  |
| --- | --- | --- | --- | --- | --- | --- | --- |
|  |  | Pooled | Original | Ct Difference | Pooled | Original | Ct Difference |
| 1 | NE-254 | 35.49 | 32.18 | 3.31 | 35.13 | 33.22 | 1.91 |
| 2 | NE-284 | 35.27 | 33.33 | 1.94 | 36.5 | 34.23 | 2.27 |
| 3 | NE-287 | 33.9 | 30.25 | 3.65 | 33.92 | 31.69 | 2.23 |
| 4 | NE-327 | 33.24 | 30.44 | 2.8 | 32.56 | 30.52 | 2.04 |
| 5 | NE-379 | 29.23 | 24.2 | 5.03 | 28.52 | 24.08 | 4.44 |
| 6 | NE-393 | 35.5 | 34 | 1.5 | 36.72 | 35.33 | 1.39 |
| 7 | NE-479 | 20.57 | 18.23 | 2.34 | 19.18 | 17.33 | 1.85 |
| 8 | NE-464 | 23.93 | 21.2 | 2.73 | 23.07 | 20.95 | 2.12 |
| 9 | NE-616 | 33.8 | 31.17 | 2.63 | 33.61 | 31.27 | 2.34 |
| 10 | NE-784 | 33.84 | 32.79 | 1.05 | 34.17 | 32.34 | 1.83 |
| 11 | NE-796 | 24.63 | 23.07 | 1.56 | 25.04 | 23.62 | 1.42 |
| 12 | NE-822 | 31.57 | 29.71 | 1.86 | 33.29 | 30.06 | 3.23 |
| 13 | NE-863 | 33.4 | 29.69 | 3.71 | 32.1 | 30.64 | 1.46 |
| 14 | NE-875 | 23.12 | 21.08 | 2.04 | 23.79 | 21.32 | 2.47 |
| 15 | NE-886 | 22.65 | 19.34 | 3.31 | 22.01 | 20.33 | 1.68 |
| 16 | NE-892 | 24.65 | 21.4 | 3.25 | 24.68 | 22.83 | 1.85 |
| 17 | NE-901 | 32.48 | 30.19 | 2.29 | 32.92 | 32.92 | 0 |
| 18 | NE-907 | 27.7 | 25.01 | 2.69 | 27.91 | 26.34 | 1.57 |
| 19 | NE-912 | 27.91 | 24.55 | 3.36 | 28.9 | 25.06 | 3.84 |
| 20 | NE-914 | 33.71 | 30.66 | 3.05 | 33.72 | 31.66 | 2.06 |
| 21 | NE-1319 | 36.13 | 32.31 | 3.82 | 36.81 | 33.4 | 3.41 |
| 22 | NE-1437 | 36.04 | 34.72 | 1.32 | 37.57 | 33.12 | 4.45 |
| 23 | NE-1421 | 37.97 | 35.46 | 2.51 | 39.10 | 36.20 | 2.9 |
| 24 | NE-1631 | 39.86 | 36.74 | 3.12 | 39.97 | 37.09 | 2.88 |
| 25 | NE-1683 | 35.52 | 33.63 | 1.89 | 37.78 | 37.43 | 0.35 |

Abbreviations: Ct, cycle threshold
<sup>a</sup>: The extraction platforms included both automated and manual procedures.
<sup>b</sup>: The N1 and N2 targets were used to detect SARS-CoV-2 during the PCR assay.

Pools were created using 50 µl from a confirmed NP positive patient specimen added to 50 µl from each of 4 negative NP patient samples for a final volume of 250 µl. Nucleic acid (NA) extraction was performed on each pool using either the QIAGEN EZ1 Virus Mini Kit v2.0 (QIAGEN, Germantown, MD) or the QIAGEN manual extraction kit according to manufacturer’s instructions. Real-time RT-PCR was performed on the extracted NA using the CDC Diagnostic Panel following the manufacturer’s instructions. The results showed that all 25-pooled specimens were positive within a range of 0 Ct to 5.03 Ct difference from the original samples (Table 1). To examine this approach in a clinical situation, 60 specimens from individuals at risk for COVID-19 as determined by the public health department were separated randomly into 12 pools, which were processed as described. Two of the pools were characterized as “2019-nCoV detected” by the assay. All individual specimens within each of the two identified pools were re-tested with two positive samples identified for an overall positive rate of 3.3%. The total reactions used were 22 for an overall conservation of 38 extraction kits and 38 amplification reagents.

Group testing of pooled samples has been successfully employed by the blood procurement and infectious disease testing for many years^5^. The strategy became effective due to the development of highly sensitive molecular based assays and several studies reported on statistical measures to determine appropriate parameters for use^6^. This study examined whether pooling was feasible using an EUA SARS CoV-2 assay in a public health setting where the desire to test large numbers of individuals has been impacted by the scarcity of key resources. The predictive algorithm indicated a pooling ratio of 1 to 5 was expected to retain accuracy of the test and result in greater efficiency of test resources. Results of this study indicated that all positive samples by the non-pooled method were detected in pools with four other negative samples.

The practical application of this process was confirmed with 60 samples from the community resulting in the saving of reagents and personnel time that could expand testing to an additional 38 samples. Assuming a consistent positivity rate, this strategy would expand testing by 133%. Table 2 summarizes the impact of different positive test rates on the overall efficiency of test resources.

**Table 2.** Comparison of optimal pool size and prevalence rates on test efficiency ^a^.

| Prevalence rate<br>(%) | Optimal specimen<br>pool size | Reduction in the expected<br>number of tests (%) | Expected increase in testing<br>efficiency (%) |
| --- | --- | --- | --- |
| 1 | 11 | 80 | 400 |
| 3 | 6 | 67 | 200 |
| 5 | 5 | 57 | 133 |
| 7 | 4 | 50 | 100 |
| 10 | 4 | 41 | 69 |
| 15 | 3 | 28 | 39 |

a. The Shiny application for pooled testing available at https://www.chrisbilder.com/shiny was used for calculations.

During a rapidly changing epidemic, testing strategies will need to adapt to potential increases in the positive test rate. Group testing of pooled specimens also requires the use of highly sensitive assays to avoid missing low positive samples. Therefore, strategies must be employed to closely monitor the use of pooling as the positive rate of test specimens increases in an outbreak of disease. Additionally, the impact of different extraction methods on the recovery of RNA and overall test sensitivity need to be evaluated. Therefore, laboratories must perform their own validation pool studies for kits used for each RNA extraction and amplification based on the prevalence rate of COVID-19 in their own region. Finally, this study showed that pooling is an effective approach to expand the impact of limited test resources and reagents during specific stages of an infectious disease outbreak.

## Data Availability

All data is available in the main text, tables and the figure.

## Acknowledgement

The authors thank the staff of the Biology Section within the Nebraska Public Health Laboratory for processing of the specimens and performance of the assay. According to the University of Nebraska Medical Center Institutional Review Board, this study was deemed exempt because it was conducted as a part of a diagnostic testing study.

The FDA reviewed the procedure as part of a EUA approved diagnostic procedure. All authors declare no conflict of interest.

## References

1. Del Rio C, Malani PN. COVID-19-New Insights on a Rapidly Changing Epidemic. JAMA. 2020 Feb 28. doi: 10.1001/jama.2020.3072.

2. Sharfstein JM, Becker SJ, Mello MM. Diagnostic Testing for the Novel Coronavirus. JAMA. 2020 Mar 9. doi: 10.1001/jama.2020.3864.

3. Iwen PC, Stiles KL, Pentella MA. Safety Considerations in the Laboratory Testing of Specimens Suspected or Known to Contain the Severe Acute Respiratory Syndrome Coronavirus 2 (SARS-CoV-2). Am J Clin Pathol. 2020. Mar 19. pii: aqaa047. doi: 10.1093/ajcp/aqaa047.

4. Van TT, Miller J, Warshauer DM, et al. Pooling nasopharyngeal/throat swab specimens to increase testing capacity for influenza viruses by PCR. J Clin Microbiol. 2012;50(3):891–896. doi: 10.1128/JCM.05631-11

5. Dwyre DM, Fernando LP, Holland PV. Hepatitis B, hepatitis C and HIV transfusion-transmitted infections in the 21st century. Vox Sang. 2011;100(1):92–98. doi: 10.1111/j.1423-0410.2010.01426.x.

6. Hitt BD, Bilder CR, Tebbs JM et al. The objective function controversy for group testing: Much ado about nothing? Stat Med. 2019;38(24):4912–4923. doi: 10.1002/sim.8341.

